# Validation of HemoCue method and development of a revised hemoglobin cut-off to detect anemia in children aged 6-24-months

**DOI:** 10.64898/2026.03.22.26349043

**Authors:** Mustafa Mahfuz, Ar-Rafi Khan, S. M. Tafsir Hasan, Md. Shabab Hossain, A.H.M Rezwan, Mehjabin Tishan Mahfuz, Md. Ashraful Alam, Tahmeed Ahmed

## Abstract

**Background:** Accurate hemoglobin (Hb) measurement is essential for determining anemia prevalence at the population level. The HemoCue method, commonly used in resource poor settings, has been shown to overestimate Hb levels compared to the direct cyanmethemoglobin (DCM) method, leading to an underestimation of anemia. This study aimed to validate the HemoCue method against the DCM method and establish an optimal cut-off value for diagnosing anemia in children under two.

**Methods:** Date were collected from children under two years of age as part of the Etiology, Risk Factors, and Interactions of Enteric Infections and Malnutrition and the Consequences for Child Health and Development study in Bangladesh, conducted in Mirpur, Dhaka, from February 2010 to February 2017. Using HemoCue 201 instrument and DCM method, Hb levels was measured in 846 venous blood samples collected from 589 children. Youden’s Index was used to identify the optimal cut-off for hemoglobin. While it provided a strong threshold, we prioritized achieving at least 70% sensitivity and specificity to confirm balanced diagnostic performance.

**Results:** The median (IQR) Hb concentrations estimated by HemoCue and DCM were 11.6 (10.5, 12.5) g/dL and 10.8 (10.1, 11.5) g/dL, respectively. Using a cut-off of <10.5 g/dL for anemia, the prevalence was significantly higher with the DCM method (35.2% vs. 23.2%). Considering DCM as the gold standard, the sensitivity and specificity of the HemoCue method were found to be 93% and 53%, respectively. In the test/train analysis, the newly identified cut-off of 11.00 g/dL, indicated an anemia prevalence of 34.2% in the testing dataset, with a sensitivity of 84% and specificity of 69%.

**Conclusion:** This study underscores the discrepancies between the HemoCue and DCM methods for measuring Hb levels. By proposing and validating a new cut-off value, we provide a more accurate means of diagnosing anemia in children under two.

## Introduction

Anemia is a major public health issue, contributing to chronic infections and nonspecific diseases [1,2]. Globally, around 1.8 billion people are affected by anemia, with a prevalence of 39.8% in children under five years old [3–6]. The consequences of anemia include impaired mental and physical growth, altered social behavior, and reduced cognitive performance [3,7,8]. These impacts can extend to poor educational outcomes and, over time, harm a country’s economic growth, with an estimated cost ranging from $7,000 to $29,000 USD per case and up to 4% of GDP loss due to iron deficiency, the leading cause of anemia [9–12].

The World Health Organization (WHO) defines anemia in children under two as a hemoglobin (Hb) concentration below 10.5 g/dL [13]. Hb concentration is the most common indicator for anemia, and the cyanmethemoglobin method is the gold standard recommended by the International Committee for Standardization in Hematology [14–16]. Although automated hematology analyzers are accurate, they are expensive and impractical for large-scale or field-based studies [17]. The HemoCue photometric method offers a more portable, cost-effective alternative, providing digital Hb readings within 10 seconds using a small blood sample without requiring electricity or refrigeration [16,18,19]. This device is portable, user-friendly, and time-efficient, making it ideal for fieldwork [16,18,20]. However, several studies have indicated that the HemoCue system may overestimate mean Hb values when compared to the cyanmethemoglobin method, in both capillary and venous blood samples [14,21,22]. This discrepancy in Hb estimation may lead to an underestimation of anemia prevalence at the population level.

This study, conducted in Dhaka’s urban slums, compares the sensitivity and specificity of the HemoCue method against the cyanmethemoglobin method using venous blood samples in children under two. The paper aims to validate the HemoCue method in comparison to the cyanmethemoglobin standard and proposes a revised hemoglobin cut-off value for detecting anemia in this population.

## Methods

This study utilized hemoglobin (Hb) data from children under two years of age, collected as part of the Etiology, Risk Factors, and Interactions of Enteric Infections and Malnutrition and the Consequences for Child Health and Development (MAL-ED) study in Bangladesh (PR-2008-020) (ClinicalTrials.gov Identifier: NCT02441426), conducted in Mirpur, Dhaka, from February 2010 to February 2017. Data for present study was collected between 04 October 2010 and 05 June 2013. The primary objective of the study was to explore the relationships between various risk factors for malnutrition, gut physiology, microbial exposure, infections, diet, nutrition, and their impacts on physical and cognitive development as well as immune responses [23]. Comprehensive descriptions of the MAL-ED study and its Bangladesh site have been published elsewhere [23].

The Bangladesh MAL-ED study comprised two main components: (i) a birth cohort study and (ii) a case-control study. In the birth cohort component, children were enrolled within 17 days of birth, following clearly defined inclusion and exclusion criteria, and were followed until five years of age. Blood samples were collected at 7, 15, 24, 36, and 60 months. In contrast, the case-control component was a prospective community-based intervention study that enrolled two groups of children aged 6 to 24 months and followed them for five months. The case group consisted of severely to moderately underweight children (weight-for-age z-score < -2, based on the WHO 2006 growth reference), while the control group included age- and community-matched well-nourished children (WAZ > -1). In the case-control study, venous blood samples were collected at enrollment and five months after enrollment. Hemoglobin levels were measured in these samples using both the HemoCue 201 instrument and direct cyanmethemoglobin (DCM) methods [23].

### Blood Collection, Processing, and Storage

Approximately 5 mL of venous blood was collected from each of the 589 participating child using a butterfly needle system, following a standard operating procedure approved by the ethical review board. After collection, the blood samples were immediately placed in a cooler with cold packs and centrifuged within two hours. All samples were processed in laboratories across the study sites using standardized procedures. After processing, the samples were stored in –80°C freezers for subsequent laboratory analyses [24,25].

### Hemoglobin measurement

Hemoglobin concentration was measured immediately after blood collection using a HemoCue 201 instrument [26]. Additionally, the colorimetric cyanmethemoglobin method was employed to measure Hb, where total Hb in blood at an alkaline pH is rapidly converted into a cyanoderivative. The absorbance of this cyanoderivative was then measured at 540 nm [27].

### Statistical analysis plan

Data were summarized either as means and standard deviations (SD) or median (inter quartile range (IQR)), as appropriate. Anemia diagnosis was made using a cut-off point of <10.50 g/dl for Hb recommended by the WHO [13]. Hemoglobin measurements from venous blood using the DCM method were considered the gold standard for diagnosing anemia. Both the HemoCue and DCM methods were used to diagnose anemia based on venous samples. Inter-rater reliability for anemia diagnosis between the two methods was assessed using Cohen’s kappa statistic. McNemar’s test was employed for paired anemia data, and Pearson’s correlation coefficient was calculated using the pairwise method [28].

Agreement between the DCM and HemoCue methods was evaluated using the Bland-Altman plot [29]. An initial linear regression analysis was conducted to examine the relationship between the two methods and to estimate a potential correction factor for improving measurement accuracy. However, the assumption of linearity was not satisfied. Then, a quadratic model was fitted to enhance model fit and address residual patterns, but the linearity assumption doesn’t hold. Therefore, no correction factor can be recommended.

Sensitivity, specificity, positive predictive value (PPV), and negative predictive value (NPV) were estimated for diagnosing anemia using the HemoCue method. A receiver operating characteristic (ROC) curve analysis was performed to assess Hb levels measured by the HemoCue method in relation to anemia with the area under curve calculated to assess classification performance [30]. Youden’s Index was used to identify the optimal cut-off point for Hb measured by the HemoCue method [31]. To ensure a clinically balanced diagnostic cut-off point, we did not rely solely on Youden’s Index. Instead, identifying a cut-off that achieves at least 70% sensitivity and specificity. Nearby alternative thresholds were also assessed to determine whether any cut-off points provided meaningful improvements in terms of statistical robustness and biological relevance.

To ensure adequate precision, particularly sensitivity and specificity, we performed the required sample size calculation as part of power analysis. With a 5% significance level and precision of 5% for both sensitivity and specificity, the following formulas were used to determine the required sample size:

Sensitivity:

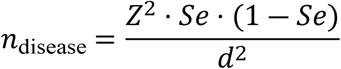

Specificity:

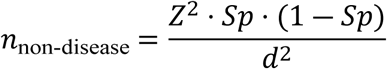

Where:

𝑍 = Z-score for the 95% confidence level

𝑆𝑒 = Expected sensitivity

𝑆𝑝 = Expected specificity

𝑑 = Desired precision 5%

The sample size was calculated to estimate the respective proportions. The total sample size was adjusted to account for disease prevalence. This approach ensures that the study is sufficiently powered and provide reliable estimates of both sensitivity and specificity.

The dataset was randomly divided into training (70%) and testing (30%) sets for validation. All statistical analyses were conducted using R software (version 4.5.2).

### Ethics

The Ethical Review Committee (ERC) of the International Centre for Diarrhoeal Diseases and Research, Bangladesh (icddr,b), has approved the study, with the protocol number PR-2008-020. Before participation in the study, one of the parents or legal guardians of the participants provided written informed consent, affirming their willingness to participate after being fully informed about the study’s nature, purpose, risks, and benefits.

## Results

A total of 265 healthy newborns were enrolled in the birth cohort study within 17 days of birth, following strict inclusion criteria. Plasma samples were collected at 7 and 15 months. Hemoglobin was measured using both the DCM and HemoCue methods with 173 participants assessed at 7 months and 165 participants assessed at 15 months. In the case-control component of the MAL-ED study, 500 cases and 480 controls were enrolled. Hemoglobin data from both DCM and HemoCue methods were available for 87 cases and 178 controls at baseline, and for 99 cases and 144 controls at the endline (5 months post-enrollment). Overall, 846 Hb assessments from venous blood were obtained using both methods (Fig 1). The median (IQR) age of children at the time of blood collection was 13.00 (7.00, 15.00) months, and half were girl.

**Fig 1.**
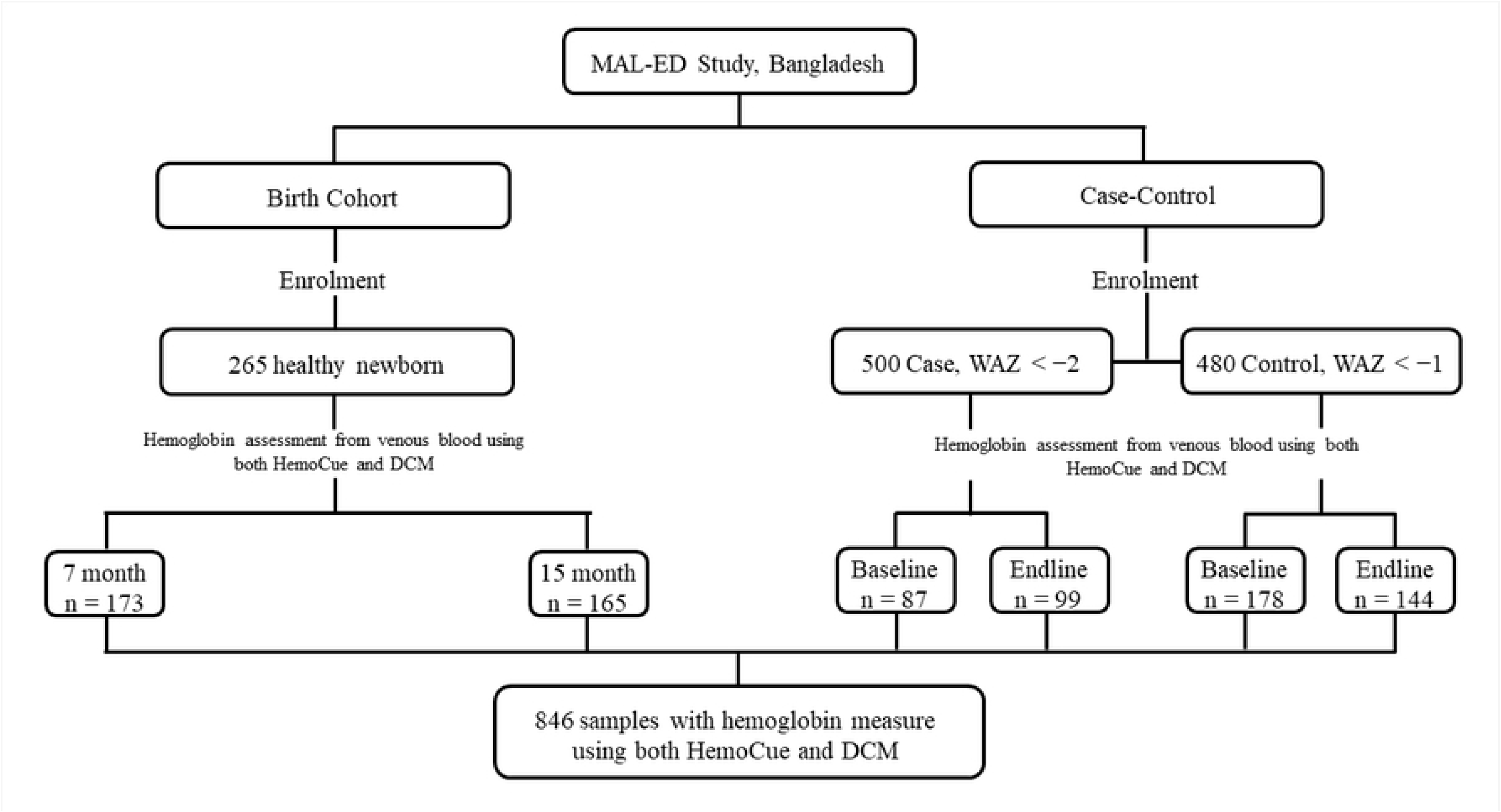
Sample selection flow chart from different cohorts used in the analysis.

In the MaL-ED birth cohort study, 265 healthy newborns were enrolled within 17 days of birth. Plasma samples were collected at 7 and 15 months. Hemoglobin levels were measured using both the direct cyanmethemoglobin (DCM) and HemoCue methods for 173 and 165 participants, respectively. In the case-control portion of the study, there were 500 cases and 480 controls. Hemoglobin data from both DCM and HemoCue methods were accessible for 87 cases and 178 controls at baseline, as well as for 99 cases and 144 controls at the endline. In total, 846 Hb assessments from venous blood were performed using both methods.

In terms of growth metrics, the mean (SD) length-for-age z-score (LAZ) was -2.70 (0.87) in the case group, -0.99 (0.84) in the control group, and -1.53 (0.99) in the birth cohort group. The mean weight-for-age z-score (WAZ) was -2.64 (0.68) in the case group and -0.50 (0.68) in the control group, with the cohort group having a mean WAZ of -1.13 (1.05). The mean weight-for-length z-score (WLZ) was -0.40 (1.06) for the birth cohort, -1.63 (0.83) for the case group, and 0.03 (0.77) for the control group (Table 1).

**Table 1.** Characteristics of participants in different cohorts including average anthropometry and hemoglobin levels.

| Indicator | Case | Control | Cohort | Overall |
| --- | --- | --- | --- | --- |
| n | 148 | 230 | 211 | 589 |
| Number of samples | 186 | 322 | 338 | 846 |
| Age (month), Median (IQR) | 14.00 (11.00, 18.00) | 13.00 (9.00, 17.00) | 8.00 (7.00, 15.00) | 13.00 (7.00, 15.00) |
| Sex, n (%) (Girl) | 96 (51.6%) | 155 (48.1%) | 169 (50.0%) | 420 (49.6%) |
| Length-for-age z score, Mean (SD) | -2.70 (0.87) | -0.99 (0.84) | -1.53 (0.99) |  |
| Weight-for-age z score, Mean (SD) | -2.64 (0.68) | -0.50 (0.68) | -1.13 (1.05) |  |
| Weight-for-length z score, Mean (SD) | -1.63 (0.82) | 0.03 (0.77) | -0.40 (1.06) |  |
| Hemoglobin DCM g/dl, Median (IQR) | 10.80 (10.30, 11.50) | 11.00 (10.30, 11.80) | 10.50 (9.80, 11.30) | 10.80 (10.10, 11.50) |
| Hemoglobin HemoCue g/dl, Median (IQR) | 11.60 (10.60, 12.50) | 12.10 (11.00, 12.80) | 11.20 (10.30, 12.10) | 11.60 (10.50, 12.50) |
| Anemia (DCM), n (%) (Yes) | 59 (31.7%) | 92 (28.6%) | 147 (43.5%) | 298 (35.2%) |
| Anemia (HemoCue), n (%) (Yes) | 42 (22.6%) | 50 (15.5%) | 104 (30.8%) | 196 (23.2%) |

Abbreviations: IQR- Inter quartile range, SD- Standard Deviation, DCM- direct cyanmethemoglobin method
Note: Number of samples were considering all timepoint

In the case group, median (IQR) Hb measured using the DCM method was 10.80 (10.30, 11.50) g/dL, compared to 11.00 (10.30, 11.80) g/dL in the control group, and 10.50 (9.80, 11.30) g/dL in the birth cohort group. With a cut-off point >10.5 g/dl for anemia, anemia prevalence was 31.7% of the case group, 28.6% of the control group, and 43.5% of the birth cohort group. Using the HemoCue method, the mean Hb was 11.60 (10.60, 12.50) g/dL for the case group, 12.10 (11.00, 12.80) g/dL for the control group, and 11.20 (10.30, 12.10) g/dL for the birth cohort. Anemia was diagnosed in 22.6% in the case group, 15.5% in the control group, and 30.8% in the birth cohort group (Table 1).

By pooling data from all groups and repeated measurements, the median (IQR) Hb concentration measured by the DCM method was 10.8 (10.1, 11.5) g/dL, and 35.2% of children were classified as anemic. Using the HemoCue method, the median (IQR) Hb concentration was 11.6 (10.5, 12.5) g/dL, with 23.2% of children diagnosed with anemia (S1 Table). We observed a strong correlation between the DCM and HemoCue methods, with a Pearson’s correlation coefficient of 0.72. McNemar’s test revealed a statistically significant difference between the methods (p < 0.001), and the Kappa statistic (79.20% agreement, kappa = 0.51, p < 0.001) indicated moderate agreement between the two methods.

To further assess agreement between the DCM and HemoCue methods, we constructed a Bland-Altman plot, where the difference and average of the two methods were calculated for each observation. The plot showed that the average difference (i.e., bias) was close to zero, and more than 95% of the data fell between the lower and upper limits of agreement (Fig 2, S1 Text). A polynomial regression of degree 2 (quadratic model) was performed to estimate a correction factor to improve measurement accuracy (S1 Fig), but linearity could not be established. We assessed linearity by stratifying Hb measured from DCM method to its nearest round number and observed a significant dispersion in Hb measurements obtained using the HemoCue method (S2 Fig). Although the agreement between the DCM and HemoCue methods was moderate to strong, our data did not allow us to propose a correction factor.

**Fig 2.**
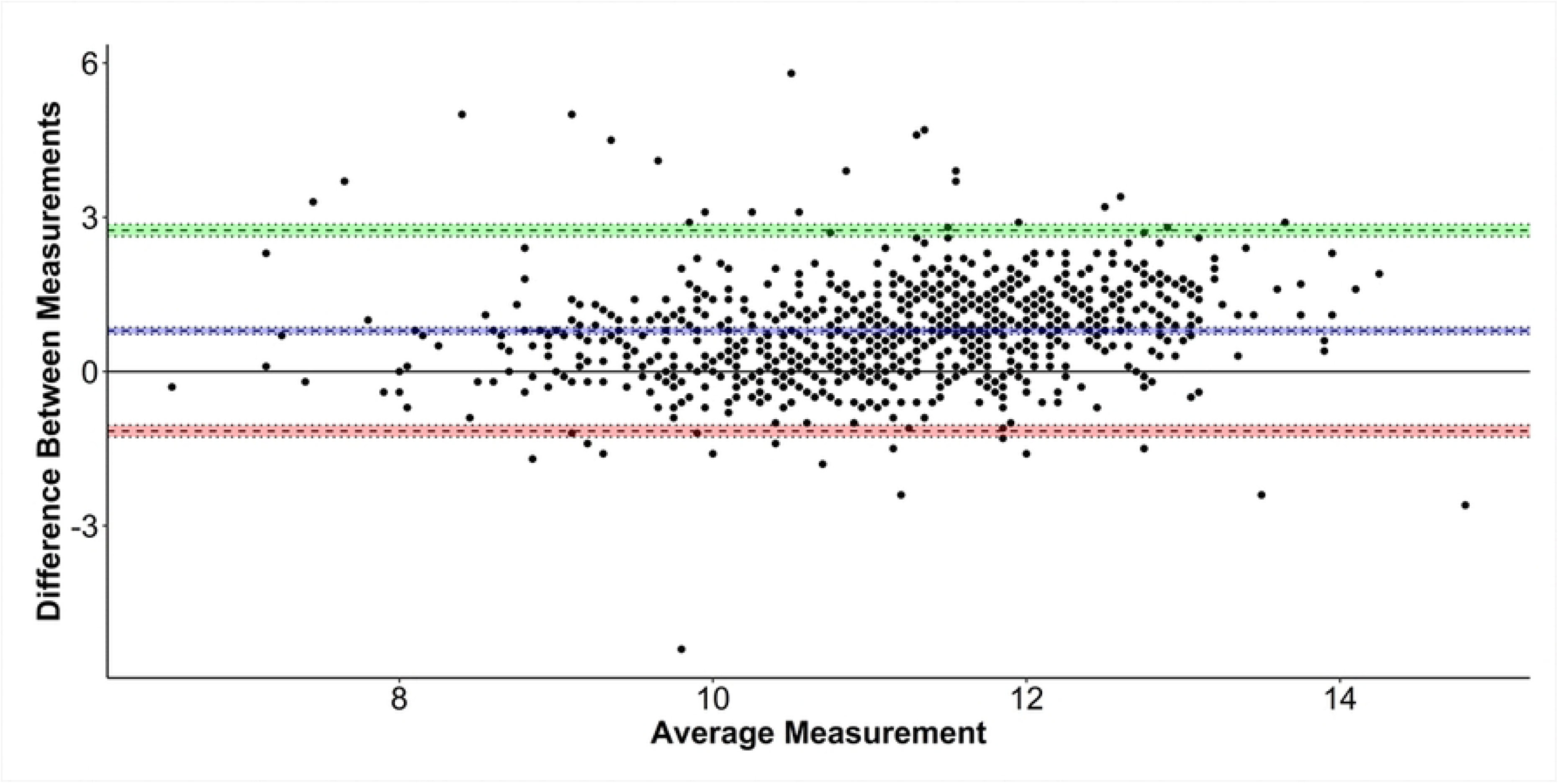
Bland-Altman plot for comparing DCM and HemoCue method. Bland–Altman plot for comparing DCM and HemoCue method agreement analysis (n = 846). Limits of Agreement are shown as dotted black lines with 95% confidence intervals (between lime and rosy-brown color areas), bias (as dotted black line) with 95% confidence interval (blue area).

We then developed a receiver operating characteristic (ROC) curve (Fig 3) using a Hb cut-off point of 10.50 g/dL to diagnose anemia (Table 2). The area under the ROC curve (AUC) was 86.55%, with an accuracy of 79.20%, sensitivity of 93.25%, and specificity of 53.36%. The positive predictive value (PPV) was 78.62%, and the negative predictive value (NPV) was 81.12%. Of the total samples, 159 were true positives, 511 were true negatives, 139 were false negatives, and 37 were false positives (Table 3). The optimal Hb cut-off point of 10.9 g/dL was identified based on Youden’s index (Fig 3A).

**Fig 3:**
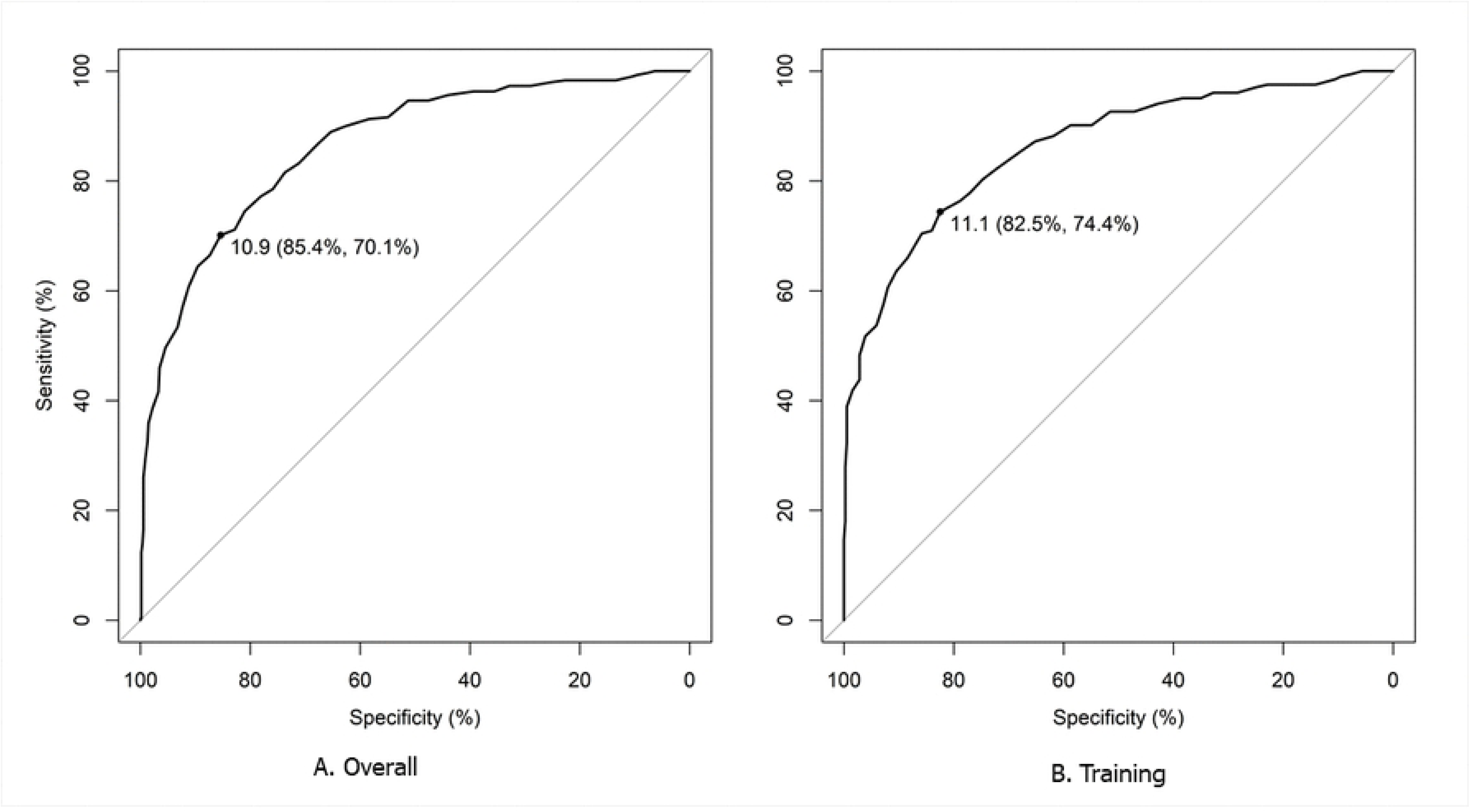
ROC curve with Youden’s index. Youden’s index is the value that maximizes the sensitivity and specificity. (A) The optimal hemoglobin cut-off point of 10.9 g/dL was identified based on Youden’s index. (B) The cut-off point of 11.1 g/dL was confirmed in the training set using Youden’s index.

**Table 2:** Summary results of ROC curves from all hemoglobin data and training data.

| ROC analysis | All Hb data | Training data |
| --- | --- | --- |
| n | 846 | 591 |
| Area under ROC curve | 86.55% | 86.26% |
| Accuracy | 79.20% | 80.20% |
| 95% CI | 76.30%, 81.88% | 76.76%, 83.34% |
| Sensitivity | 93.25% | 94.07% |
| Specificity | 53.36% | 53.69% |
| Positive predictive value | 78.62% | 79.52% |
| Negative predictive value | 81.12% | 82.58% |
| True positive | 159 | 109 |
| False negative | 139 | 94 |
| False positive | 37 | 23 |
| True negative | 511 | 365 |

Abbreviations: Receiver-operating characteristic curve (ROC)

**Table 3:** Different hemoglobin cut-off metric age 6 to 24 month.

| Metric | Cut-point<br>10.80 | Cut-point<br>10.90 | Cut-point<br>11.00 | Cut-point<br>11.10 |
| --- | --- | --- | --- | --- |
| Accuracy | 80.73% | 80.02% | 80.02% | 78.72% |
| Kappa | 0.561 | 0.552 | 0.559 | 0.537 |
| McNemar's Test (p-value) | <0.001 | 0.021 | 0.538 | 0.602 |
| Precision | 0.82 | 0.83 | 0.84 | 0.84 |
| Sensitivity (True Positive Rate) | 89.60% | 87.41% | 85.40% | 82.85% |
| Specificity (True Negative Rate) | 64.43% | 66.44% | 70.13% | 71.14% |
| Positives predicted value | 82.24% | 82.73% | 84.02% | 84.07% |
| Negatives predicted value | 77.11% | 74.16% | 72.32% | 69.28% |
| Detection Rate | 58.04% | 56.62% | 55.32% | 53.66% |
| Balanced Accuracy | 77.01% | 76.93% | 77.77% | 76.99% |

A threshold value of 11.00 achieved the most favorable trade-off between sensitivity (85.4%) and specificity (70.1%), resulting in the greatest balanced accuracy (77.78%) across the examined cut-offs. At this point, precision remained high (84.02%), and the kappa statistic (0.559) reflected moderate concordance with the reference standard. In addition, McNemar’s test did not indicate a statistically significant disagreement, further supporting the stability and reliability of this threshold (Table 3).

Next, we validated the new cut-off of 11.00 g/dL through a split-sample analysis. Seventy percent of the data were randomly allocated to a training set and 30% to a test set. In the training set, the AUC was 86.26%, with a sensitivity of 94.07% and a specificity of 53.69% (Table 2).

For the test set, ROC analysis showed an AUC of 86.77%, with a sensitivity of 91.25% and a specificity of 52.63%. After applying the new cut-off of 11.00 g/dL to the test set, the specificity has improved to 69.47% (Table 4). Using this new cut-off, 34.2% of children were classified as anemic, compared to 23.2% with the current cut-off of 10.5 g/dL (Table 5).

**Table 4:** Summary results of ROC curve for test dataset using both the cutoff points.

|  | Cut-off Hb 10.5 g/dL | Cut-off 11.0 g/dL |
| --- | --- | --- |
| The area under ROC curve | 86.77% | 100.0% |
| Accuracy | 76.86% | 78.82% |
| 95% CI | 71.19%, 81.90% | 73.29%, 83.67% |
| Sensitivity | 91.25% | 84.38% |
| Specificity | 52.63% | 69.47% |
| Positive predictive value | 76.44% | 82.32% |
| Negative predictive value | 78.12% | 72.53% |
| True positive | 50 | 66 |
| False-negative | 45 | 29 |
| False positive | 14 | 25 |
| True negative | 146 | 135 |

Abbreviations: Receiver-operating characteristic curve (ROC)

**Table 5:**
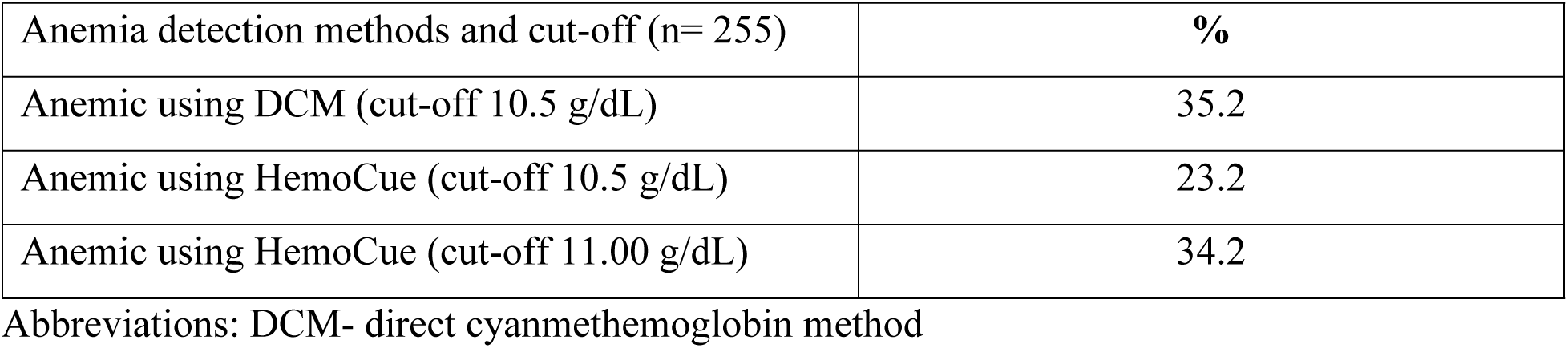
Anemia detection in test dataset with new and old cut-offs.

| Anemia detection methods and cut-off (n= 255) | % |
| --- | --- |
| Anemic using DCM (cut-off 10.5 g/dL) | 35.2 |
| Anemic using HemoCue (cut-off 10.5 g/dL) | 23.2 |
| Anemic using HemoCue (cut-off 11.00 g/dL) | 34.2 |

Abbreviations: DCM- direct cyanmethemoglobin method

## Discussion

Identification of anemia at the population level remains a challenge due to limited diagnostic infrastructure. Children under two years old are vulnerable to anemia and the accurate measurement of Hb is limited in resource poor settings [10]. In this study, we compared the HemoCue method to the gold standard DCM method. Our findings revealed significant discrepancies between the two methods, which can influence the reported prevalence of anemia and subsequent public health decisions.

We compared the Hb levels of the children age under two enrolled in the MAL-ED study, using both the DCM and HemoCue methods. The median (IQR) Hb concentration measured by the DCM method was 10.8 (10.1, 11.5) g/dL, with 35.2% of children classified as anemic. HemoCue method reported a higher median Hb level of 11.6 (10.5, 12.5) g/dL, with 23.2% of children diagnosed as anemic. Our results showed a 34% underestimation of anemia prevalence in the community when using HemoCue. There is a moderate Pearson’s correlation (r = 0.72) between DCM and HemoCue method. Statistically significant difference was observed in McNemar’s test (p < 0.001) highlights the challenge of using HemoCue for accurate anemia diagnosis. Cohen’s kappa (k = 0.51, p < 0.001) indicates moderate agreement between the two methods. These differences suggest that anemia prevalence might be significantly underestimated when relying solely on the HemoCue method, a finding consistent with previous studies [14,22,32]. The Bland-Altman plot showed a good agreement between the two methods, with over 95% of observations falling within the limits of agreement. This suggests, though the HemoCue method tends to overestimate Hb levels, the overall difference remains within an acceptable range [33].

Although we attempted to develop a correction factor through polynomial regression, the lack of linearity prevented its development. Then we explored various methods to understand why linearity was not achieved, including stratification by age and Hb levels. When Hb values measured by the DCM method were rounded to the nearest whole number, we observed a considerable dispersion in Hb levels measured by the HemoCue method (S2 Table, S1 and 2 Figs). This wide variation in HemoCue measurements likely accounts for the failure to achieve linearity in the analysis. Despite a moderate to strong agreement between the methods, no correction factor could be proposed based on our data. Therefore, our analysis led to the identification of a new optimal cut-off value of 11.00 g/dL, to better align the HemoCue method with DCM findings. Validation using the training-test approach showed that the new cut-off improved both sensitivity and specificity, with the test dataset achieving 84.38% sensitivity and 69.47% specificity, a marked improvement over the previous cut-off of 10.50 g/dL. Applying this updated cut-off increased the anemia prevalence from 23.2% to 34.2%. This result better aligns with the DCM classification, suggesting a more accurate reflection of anemia burden in this population.

Other studies comparing the HemoCue with automated hematology analyzers, such as the XT1800i (Sysmex) and BC-3000Plus (Mindray), have similarly reported poor agreement. These discrepancies resulted in significant differences in estimated anemia prevalence [16,33]. While study in Ghana found no significant differences between the HemoCue, cyanmethemoglobin, and Sysmex KX21N methods [34].

The largest required sample size was 544, corresponding to a Hb cut-off of 11.0 g/dL. The actual sample size of our study was 846, exceeding this requirement. Therefore, the study is adequately powered. This enhances the reliability of the findings and reduces the risk of type II error.

The strength of this study is in its use of venous blood samples, which allowed us to conduct direct comparisons of the two methods under controlled conditions. This approach minimizes biological variability often introduced by differences in blood sample type (e.g., capillary, venous, or arterial), as reported in earlier studies [35–37]. A recent study suggested that accurate and precise determination of Hb requires venous blood. The use of single-drop capillary blood is not recommended due to its high measurement variability [38]. However, a study in Hong Kong reported, the capillary point of care test (POCT HemoCue) had acceptable agreement with venous Hb in the emergency department setting [39]. Furthermore, our use of samples from a well-regulated study, including repeated measurements over time, and regular quality controls for Hemocue analyzers, enabled a robust comparison and validation of the new cut-off value. The train-test validation process also enhanced the reliability of our findings, ensuring that the new cut-off is not limited to a specific sample but generalizable across similar populations [40].

Despite these strengths, our study has a few limitations. While venous blood sampling ensures greater accuracy, it is not the standard collection method in field settings where HemoCue is often used, particularly in resource-poor areas. The validity of our findings for capillary blood samples, commonly used in community-based anemia assessments, remains unverified. Previous research has found significant variability in Hb levels depending on the site of blood collection. Future studies should therefore examine whether the proposed cut-off holds. Our study focused exclusively on children aged under two years of age in Bangladesh. This cohort provides valuable insights into the performance of the HemoCue method in a low-resource setting, the findings may not be generalizable to other regions or populations. Factors such as altitude, nutritional status, and the prevalence of genetic hemoglobinopathies could influence Hb levels and affect the applicability of this cut-off [41–43]. Validation studies across diverse geographic and demographic settings are therefore essential to ensure broader applicability. While we identified a new cut-off value to improve anemia classification using HemoCue, the inherent limitations of the device, such as its tendency to overestimate Hb persist. Efforts to improve the accuracy of portable hemoglobinometers like HemoCue should continue, as they are indispensable in large-scale surveys and fieldwork where access to laboratory-based methods is not feasible. Though these analyses incorporated repeated measures for the same children may impact the assumption of independence, we do not anticipate this to significantly affect the results or compromise their interpretability. While we identified a new cut-off value to improve anemia classification using HemoCue, the inherent limitations of the device, such as its tendency to overestimate Hb persist. Efforts to improve the accuracy of portable hemoglobinometers like HemoCue should continue. Efforts to improve the accuracy of portable hemoglobinometers like HemoCue should continue, as they are indispensable in large-scale surveys and fieldwork where access to laboratory-based methods are not feasible. While our analyses included repeated measurements from the same children, which may challenge the assumption of independence, we do not believe this significantly affects the robustness or interpretability of our findings.

Adopting a corrected cut-off of 11.00 g/dL for HemoCue use in low-resource settings would directly address the issue of systematic Hb overestimation and improve the identification of anemic children. Despite its limitations, HemoCue remains the most available option for initial anemia screening in resource-constrained environments due to its portability, ease of use, and minimal training requirements. By applying the revised cut-off value of 11.00 g/dL, health programs can more accurate estimates of anemia prevalence and reduce the risk of missing children who require treatment.

## Conclusion

Our comparison between the HemoCue and DCM methods for measuring Hb levels and detecting anemia shows that HemoCue tends to overestimate Hb levels. By proposing and validating a new cut-off value for the HemoCue method, we provide a more accurate way of diagnosing anemia in children under two. In resource poor settings, HemoCue remains a critical tool for anemia screening. It is essential to carefully consider the cut-off value to ensure accurate population-level estimates. Future research should focus on further validating this cut-off value across diverse populations and blood types. This will help ensure that the true burden of anemia is correctly identified and properly addressed in public health programs initiatives.

## Data Availability

Data related to this manuscript are available upon request, and researchers who meet the criteria for access to confidential data may contact the Research Administration of icddr,b (http://www.icddrb.org/).

## Supporting information

**S1 Text: Detailed results from Bland-Altman plot for comparing DCM and HemoCue method**

**S1 Table: Comparison of hemoglobin concentration using direct cyanmethemoglobin and HemoCue methods in children enrolled in Mal-ED studies, Bangladesh**

**S2 Table: Polynomial regression**

**S1 Figure: Polynomial regression analysis to test linearity.** To suggest a correction factor, a simple polynomial regression of degree 2 (quadratic model) was performed. However, the assumption of linearity was not satisfied. Subsequently, paired hemoglobin concentrations measured were excluded if hemoglobin concentrations using the direct cyanmethemoglobin method were less than 7 or greater than 14, approximately 2% of the data. Despite this adjustment, the linear model still failed to achieve linearity.

**S2 Figure: Scatter plot to assess of linearity by stratifying hemoglobin levels measured by DCM.** A scatter plot was created to analyze hemoglobin concentrations using both the direct cyanmethemoglobin method and the HemoCue method. The plot was stratified by rounding the direct cyanmethemoglobin values to the nearest whole number, allowing for an assessment of the distribution of hemoglobin concentrations measured by the HemoCue method.

